# Persistence of S1 Spike Protein in CD16+ Monocytes up to 245 Days in SARS-CoV-2 Negative Post COVID-19 Vaccination Individuals with Post-Acute Sequalae of COVID-19 (PASC)-Like Symptoms

**DOI:** 10.1101/2024.03.24.24304286

**Authors:** Bruce K. Patterson, Ram Yogendra, Edgar B. Francisco, Emily Long, Amruta Pise, Eric Osgood, John Bream, Mark Kreimer, Devon Jeffers, Christopher Beaty, Richard Vander Heide, Jose Guevara-Coto, Rodrigo A Mora-Rodríguez

**Affiliations:** IncellDx Inc, San Carlos, CA; Lawrence General Hospital, Lawrence, MA; Department of Medicine, St. Francis Medical Center, Trenton, NJ; Department of Emergency Medicine, Novant Health Kernersville Medical Center, Kernersville, NC; Department of Emergency Medicine, New York Presbyterian Hospital, Brooklyn, NY; Department of Anesthesiology, Stamford Hospital, CT; Department of Pathology, Marshfield Medical Center, Marshfield, Wi; Lab of Tumor Chemosensitivity, CIET / DC Lab, Faculty of Microbiology, Universidad de Costa Rica

**Author notes:** Corresponding Author: Ram Yogendra M.D. Abbreviations: PASC, post-acute sequelae of COVID-19; POVIP, post-vaccination individuals with PASC-like symptoms; NCM, non-classical monocytes; IM, intermediate monocytes; CX3CL1, C-X3-C motif chemokine ligand 1; CX3CR1, C-X3-C motif chemokine receptor 1; IL, interleukin; RANTES, regulation on activation, healthy control T-expressed and secreted; CCR, chemokine receptor; IFN, interferon; TNF, tumor necrosis factor; MIP, macrophage inflammatory protein; PBMCs, peripheral blood mononuclear cells; VEGF, vascular endothelial growth factor; LH, long hauler or PASC.

**Keywords:** COVID-19, PASC, SARS CoV-2 S1 protein, non-classical monocytes, CCR5, fractalkine

## Abstract

There have been concerning reports about people experiencing new onset persistent complications (greater than 30 days) following approved SARS-CoV-2 vaccines (BNT162b2 (Pfizer), mRNA-1273 (Moderna), Janssen (Johnson and Johnson), and ChAdOx1 nCoV-19 (AstraZeneca)). We sought to determine the immunologic abnormalities in these patients and to investigate whether the potential etiology was similar to Post-Acute Sequalae of COVID (PASC), or long COVID.

We studied 50 individuals who received one of the approved COVID-19 vaccines and who experienced new onset PASC-like symptoms along with 45 individuals post-vaccination without symptoms as controls. We performed multiplex cytokine/chemokine profiling with machine learning as well as SARS-CoV-2 S1 protein detection on CD16+ monocyte subsets using flow cytometry and mass spectrometry. We determined that post-vaccination individuals with PASC-like symptoms had similar symptoms to PASC patients. When analyzing their immune profile, Post-vaccination individuals had statistically significant elevations of sCD40L (p<0.001), CCL5 (p=0.017), IL-6 (p=0.043), and IL-8 (p=0.022). Machine learning characterized these individuals as PASC using previously developed algorithms. Of the S1 positive post-vaccination patients, we demonstrated by liquid chromatography/ mass spectrometry that these CD16+ cells from post-vaccination patients from all 4 vaccine manufacturers contained S1, S1 mutant and S2 peptide sequences. Post-COVID vaccination individuals with PASC-like symptoms exhibit markers of platelet activation and pro-inflammatory cytokine production, which may be driven by the persistence of SARS-CoV-2 S1 proteins in intermediate and non-classical monocytes. The data from this study also cannot make any inferences on epidemiology and prevalence for persistent post-COVID vaccine symptoms. Thus, further studies and research need to be done to understand the risk factors, likelihood and prevalence of these symptoms.

**Summary:** SARS CoV-2 S1 Protein in CD16+ Monocytes Post-Vaccination

## INTRODUCTION

Despite over 13 billion doses of SARS-CoV2 vaccines being administered to individuals worldwide, there have been concerning reports and publications of persistent adverse effects of the COVID-19 vaccines. These reports suggest that new-onset cardiac, vascular and neurological symptoms commenced within minutes and hours after vaccination and subsequently, were persisting for months and years^1–5^. Interestingly, many of these persistent post-vaccine symptoms are similar to symptoms associated with post-acute sequelae of COVID (PASC), or long COVID^6,7^. We recently reported that the S1 protein subunit of SARS-CoV2 was persistent in both nonclassical (CD14- CD16+) and intermediate (CD14+CD16+) monocytes several months and years after acute infection and could be a possible pathophysiological explanation for PASC^8^. Interestingly, the BNT162b2 (Pfizer), mRNA-1273 (Moderna), Janssen (Johnson and Johnson), and ChAdOx1 nCoV-19 (AstraZeneca) vaccines also deliver a synthetic S1 protein subunit directly into muscle cells to elicit an immunological response^9^.

Since there were similarities between PASC and persistent post-vaccination symptoms in individuals who never had a history of COVID infection, we sought to study if persistent S1 proteins were also present in CD16+ monocytes of both groups. One of the challenges in studying individuals with persistent post-vaccination symptoms is the lack of approved confirmatory tests to rule out previous infection or exposure to SARS-CoV2. Hence, we used a combination of patient clinical history, negative anti-nucleocapsid antibody testing and T-detect testing as a way to screen these patients.

Here, we report on 50 individuals with symptoms >1 month following vaccination (with either Pfizer, Moderna, Johnson and Johnson or AstraZeneca) that resembled the spectrum of symptoms reported in long COVID or PASC^7, 8^. We used machine learning to analyze the immune profiles in these individuals. Further, we used high parameter, single cell analysis to determine if S1 protein generated by vaccination persisted in immune cells as previously described^8^.

## MATERIALS/METHODS

### Inclusion Criteria

We included patients who received at least 1 dose of the one of the four approved COVID vaccines (Pfizer, Moderna, Johnson and Johnson, AstraZeneca) and who experienced persistent new-onset symptoms 30 days after their last inoculation.

### Exclusion Criteria

We excluded participants with a prior history of seizure disorder, migraines, neuropathy, inflammatory bowel disease, depression and anxiety disorders, chronic fatigue syndrome, Lyme disease, fibromyalgia, arthritis, COPD, asthma, chronic kidney disease, chronic heart failure (CHF), arrhythmias, bleeding disorders, and anticoagulation therapy.

Fifty participants (age range 13-65) who developed new-onset symptoms that persisted greater than 30 days after receiving either the BNT162b2 (Pfizer), mRNA-1273 (Moderna), Janssen (Johnson and Johnson), and ChAdOx1 nCoV-19 (AstraZeneca) vaccines were enrolled following written informed consent. Forty five adult participants (age range 20-70) who received one of the four approved COVID vaccines and did not report any new-onset persistent post-vaccine symptoms (greater than 30 days) were used as controls.

Vaccine inoculation dates and batch numbers were confirmed through their CDC-issued vaccine cards. Lack of prior infection was determined using patient clinical history including prior negative polymerase chain reaction (PCR) testing, anti-nucleocapsid antibody testing and T-detect testing. As demonstrated in Table 1 and Figure 1, representative patients vaccinated with all four approved vaccines were included in the study as well as the duration of the symptoms post-vaccination.

**Figure 1.**
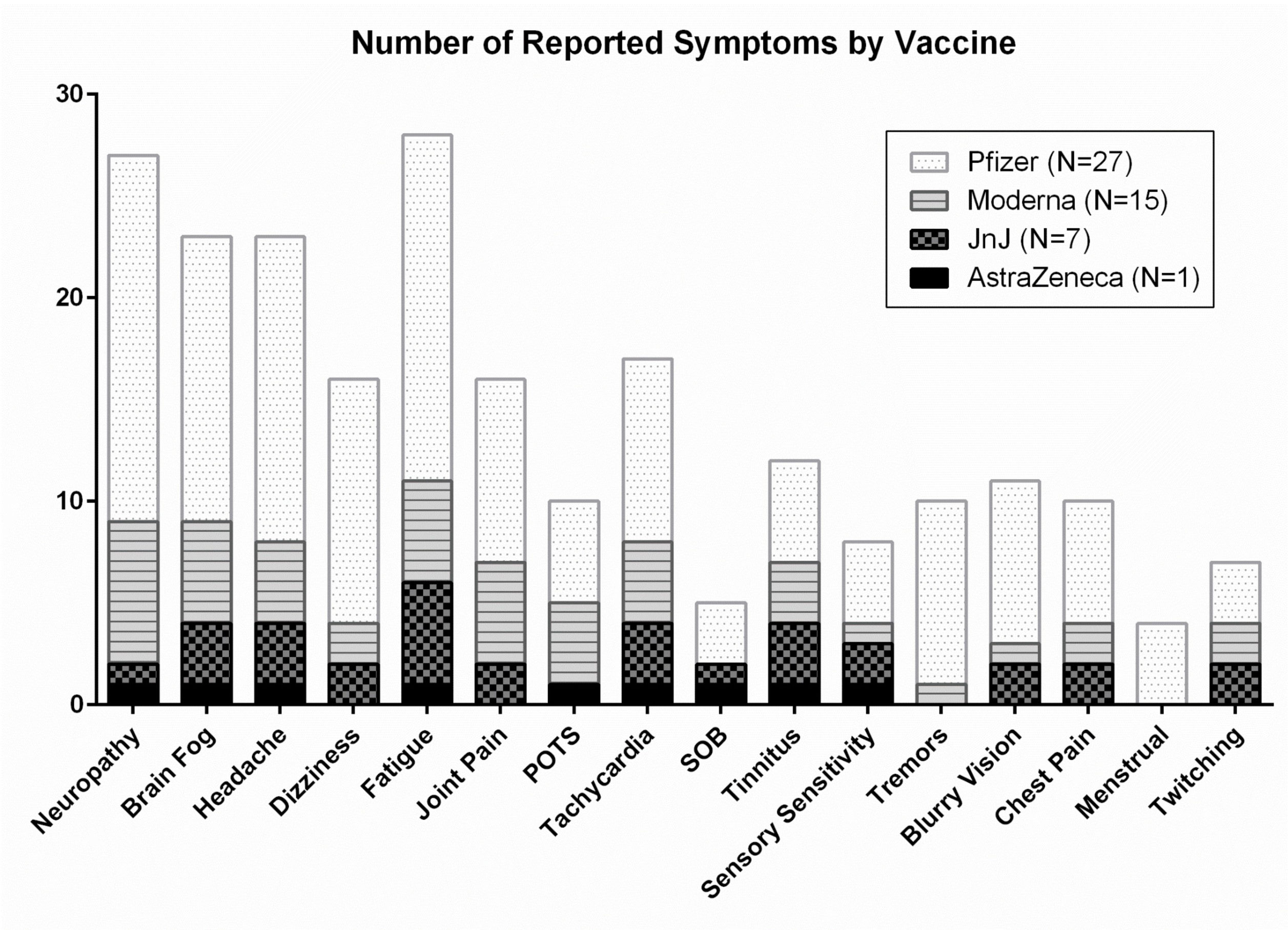
Frequency of symptoms in post-vaccination individuals with PASC-like symptoms.

**Table 1.**

|  |  | Sex | Vaccine | Days from Last<br>Vaccine to Blood<br>Draw |
| --- | --- | --- | --- | --- |
| P1 |  | F | PFE | 75 |
| P2 |  | M | MRNA | 76 |
| P3 |  | F | MRNA | 69 |
| P4 |  | F | MRNA | 128 |
| P5 |  | F | PFE | 97 |
| P6 |  | M | PFE | 45 |
| P7 |  | F | PFE | 63 |
| P8 |  | F | MRNA | 125 |
| P9 |  | F | MRNA | 142 |
| P10 |  | M | PFE | 97 |
| P11 |  | F | PFE | 97 |
| P12 |  | F | PFE | 83 |
| P13 |  | M | MRNA | 85 |
| P14 |  | F | PFE | 80 |
| P15 |  | F | PFE | 106 |
| P16 |  | M | MRNA | 79 |
| P17 |  | M | MRNA | 38 |
| P18 |  | M | MRNA | 129 |
| P19 |  | F | AZN | 245 |
| P20 |  | M | JNJ | 93 |
| P21 |  | F | PFE | 100 |
| P22 |  | F | PFE | 162 |
| P23 |  | F | MRNA | 118 |
| P24 |  | F | PFE | 149 |
| P25 |  | F | MRNA | 62 |
| P26 |  | F | PFE | 61 |
| P27 |  | F | PFE | 163 |
| P28 |  | M | JNJ | 83 |
| P29 |  | F | MRNA | 42 |
| P30 |  | F | JNJ | 168 |
| P31 |  | F | PFE | 183 |
| P32 |  | F | PFE | 82 |
| P33 |  | F | PFE | 90 |
| P34 |  | F | PFE | 70 |
| P35 |  | M | JNJ | 89 |
| P36 |  | M | PFE | 87 |
| P37 |  | F | JNJ | 91 |
| P38 |  | F | PFE | 71 |
| P39 |  | M | PFE | 88 |
| P40 |  | M | PFE | 121 |
| P41 |  | F | PFE | 63 |
| P42 |  | F | PFE | 58 |
| P43 |  | F | JNJ | 144 |
| P44 |  | F | MRNA | 39 |
| P45 |  | F | PFE | 178 |
| P46 |  | F | MRNA | 155 |
| P47 |  | F | MRNA | 179 |
| P48 |  | M | PFE | 116 |
| P49 |  | F | JNJ | 146 |
| P50 |  | F | PFE | 153 |

### High Parameter Immune Profiling/Flow Cytometry

Whole blood was collected in a 10 ml EDTA tube and a 10 ml plasma preparation tube (PPT). Peripheral blood mononuclear cells were isolated from peripheral blood using Lymphoprep density gradient (STEMCELL Technologies, Vancouver, Canada). Aliquots of 200,000 cells were frozen in media that contained 90% fetal bovine serum (HyClone, Logan, UT) and 10% dimethyl sulfoxide (Sigma-Aldrich, St. Louis, MO) and stored at -70°C for <1 week to preserve viability and run as a single batch to avoid intrarun variability. Cells were blocked with 2% BSA solution for 5 min. at RT. A cocktail containing the following antibodies and reagents was added: Brilliant Stain Buffer (BD Biosciences, San Jose, CA), True-Stain Monocyte Blocker, anti-CD19-PE-Dazzle594, antiCTLA-4 PE-Cy7, anti-CD3-APC, anti-CD16-Alexa Fluor 700 (all BioLegend, San Diego, CA), and anti-SARS-CoV-2 Spike S1 Subunit-Alexa Fluor 405 (R&D Systems, Minneapolis, MN). The following antibodies were then added: anti-CD8 BUV496, antiCD4-BUV661, anti-CD45-BUV805, anti-PD-1-BB700 (all BD Biosciences, San Jose, CA), anti-CD56-BV711 (BioLegend, San Diego, CA), and anti-CD14 BV786 (BioLegend, San Diego, CA). Cells were stained for 30 min. at RT, and then washed twice with 2%BSA. Cells were fixed for 1 hour at RT in 1 mL incellMAX (IncellDx, San Carlos, CA), and then incubated with anti- FoxP3- PE antibody (BD Biosciences, San Jose, CA) for 30 min. Cells were washed twice with 2%BSA and then acquired on a 5-laser CytoFLEX LX.

### Flow Cytometric Cell Sorting

Cryopreserved PBMCs were quick-thawed, centrifuged, and washed in 2% BSA solution in D- PBS. Cells were blocked for 5 min. in 2% BSA and then incubated at room temperature for 30 min. with Alexa Fluor® 488 Anti-CD45 antibody (IncellDx, 1/ 100 dilution), 2.5 ug of Alexa Fluor® 647 Anti-CD16 antibody (BD, Cat. # 55710), and 1 ug of PerCP/Cy5.5 Anti-human CD14 antibody (Biolegend, Cat. #325622). Cells were washed twice with 2% BSA/D-PBS, filtered, and kept on ice for the duration of the cell sort. Data was acquired on a Sony SH800, and only CD45+ cells staining positive for both CD14+ and CD16+ were sorted into test tubes with 100 uL 2% BSA solution. Sort purity of control PBMCs was confirmed to be >99% by re- analyzing sorted PBMCs using the same template and gating strategy.

### Single Cell Protein Identification

Patient cells were sorted based on phenotypic markers (as above) and frozen at -80 C. Six patient samples with positive flow cytometry signal and sufficient cell counts were chosen for LCMS confirmation. Frozen cells were lysed with the IP Lysis/Wash Buffer from the kit according to the manufacturer’s protocol. 10 ug of anti-S1 mAb were used to immunoprecipitate the S1 Spike protein from cell lysate of each patient. After overnight incubation with end-over-end rotation at 4 C and then three washes with IP Lysis/Wash Buffer, bound S1 Spike protein was eluted with the elution buffer from the kit. IP elution fractions were dried in vacuo, resuspended in 20 uL of water, pooled, and purified by Agilent 1290 UPLC Infinity II on a Discovery C8 (3cm x 2.1 mm, 5 µm, Sigma-Aldrich, room temperature) using mobile phase solvents of 0.1% trifluoroacetic acid (TFA) in water or acetonitrile. The gradient is as follows: 5- 75% acetonitrile (0.1% TFA) in 4.5 min (0.8 mL/min), with an initial hold at 5% acetonitrile (0.1% TFA) for 0.5 min (0.8 mL/ min). The purified protein was dried in vacuo and resuspended in 50 µL of 100 mM HEPES, pH 8.0 (20% Acetonitrile). 1µL of TCEP (100 mM) was added and the samples were incubated at 37°C for 30 min. 1 µL of chloroacetamide (500 mM) was added to the samples and incubated at room temperature for 30 min. 1 µL rAspN (Promega 0.5 µg/µL) and 1 µL of LysC (Pierce, 1 µg/ µL) were added and the samples incubated at 37°C for 16 h, prior to LCMS analysis.

### Liquid Chromatography/Mass Spectroscopy (LC-MS) Analysis

Digested recombinant SARS-CoV-2 Spike S1 protein was analyzed by a high accuracy mass spectrometer to generate a list of detectable peptides with retention time and accurate masses. An Agilent 1290 Infinity II high pressure liquid chromatography (HPLC) system and an AdvanceBio Peptide Mapping column (2.1 × 150 mm, 2.7 mm) were used for peptide separation prior to mass analysis. The mobile phase used for peptide separation consists of a solvent A (0.1% formic acid in H2O) and a solvent B (0.1% formic acid in 90% CH3CN). The gradient was as follows: 0–1 min, 3% B; 1– 30 min, to 40% B; 30– 33 min, to 90% B; 33-35 min, 90% B; 37-39 min, 3% B. Eluted peptides were electrosprayed using a Dual JetStream ESI source coupled with the Agilent 6550 iFunnel time-of-flight MS analyzer. Data was acquired using the MS method in 2 GHz (extended dynamic range) mode over a mass/charge range of 50– 1700 Daltons and an auto MS/MS method. Acquired data were saved in both centroid and profile mode using Agilent Masshunter Workstation B09 Data acquisition Software. The same analytical method was applied to immunoprecipitated samples from sorted patient cells except no ms/ms was acquired.

#### Machine Learning

We deployed machine learning to the acute COVID, PASC, and Post-vaccination datasets as previously described^10^ (Figure 2). Immunologic severity scores were generated using the algorithm **SC = (IL-6+sCD40L/1000 + VEGF/10 + 10*IL-10)/(IL-2 + IL-8)** and the long hauler index (PASC) was generated using the algorithm **LHI = (IFN-**γ **+ IL-2)/ CCL4-MIP-1**β.

**Figure 2.**
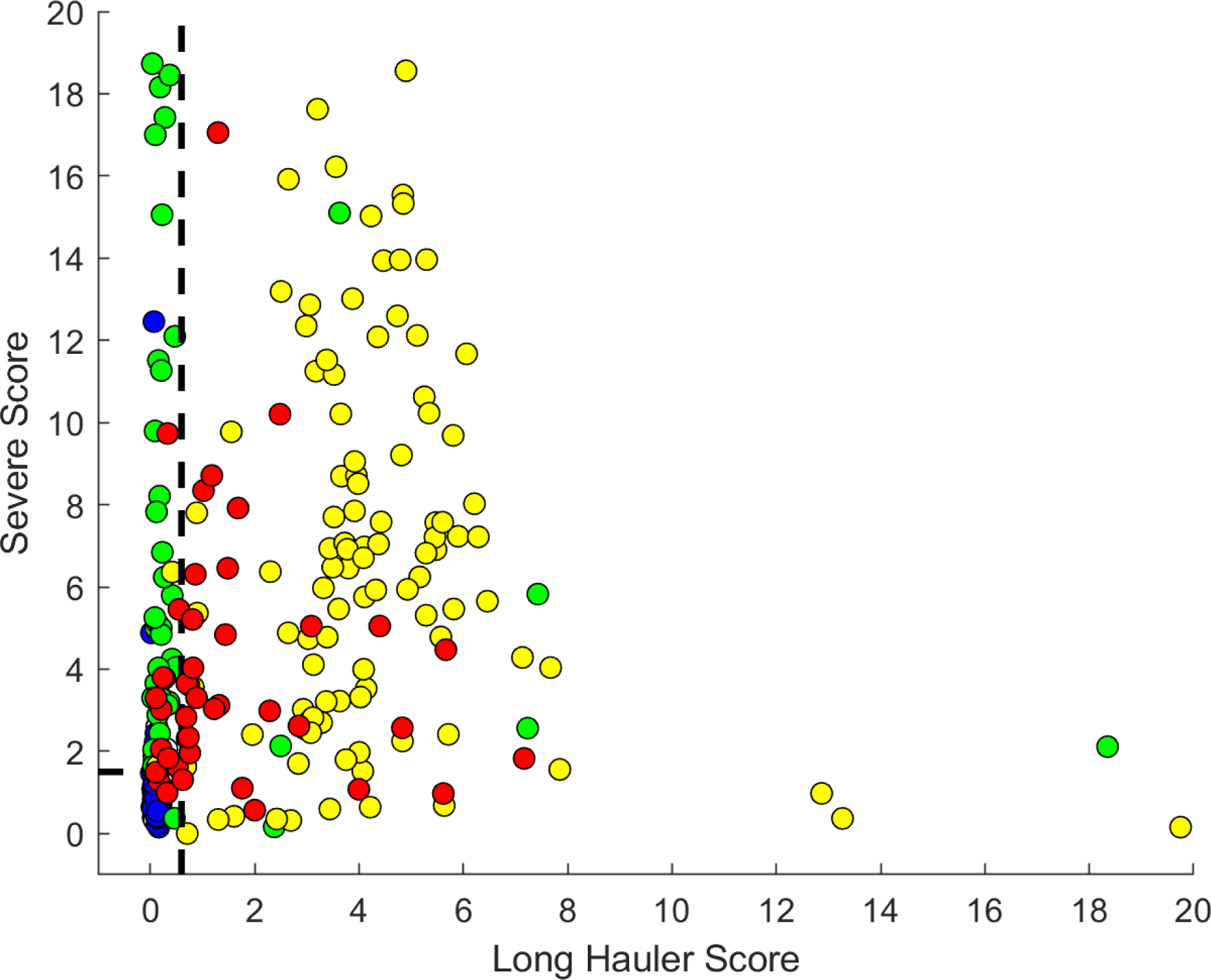
Machine learning classification of the immune profile seen in post- vaccination individuals with PASC-like symptoms (red) compared to the immune profile in individuals with PASC (yellow) due to SARS-CoV-2 infection, mild-moderate COVID infection (blue) and severe COVID (green).

## RESULTS

### Vaccine-Specific PASC-Like Symptoms

We investigated 50 individuals that exhibited PASC-like symptoms similar to those previously published ^7,10^. Table 1 shows the average age in our study was 41.8 years old with 36 females and 14 males. It also shows the average days from last inoculation with a COVID vaccine and time of blood draw for this study was 105 days. The greatest time period from inoculation to blood draw was 245 days and the shortest time period was 38 days. 27 individuals reported receiving BNT162b2 (Pfizer), 15 received mRNA-1273 (Moderna), 7 received Janssen (Johnson and Johnson), and 1 individual received ChAdOx1 nCoV-19 (AstraZeneca) vaccine.

There was variability in symptoms according to the vaccine administered, however, the predominant symptoms were fatigue (28/50), neuropathy (27/50), brain fog (23/50) and headache (23/50) (Figure 1 and Table 2). All of these symptoms were reported in all four of the vaccines administered in this cohort. Notably infrequent in the post-vaccination individuals with PASC-like symptoms compared to PASC itself were shortness of breath and loss of taste/smell.

**Table 1.**

| Neuropathy | Brain Fog | Headache | Dizziness | Fatigue | Joint Pain | POTS | Tachycardia |
| --- | --- | --- | --- | --- | --- | --- | --- |
| 27 | 23 | 23 | 16 | 28 | 16 | 10 | 17 |

| SOB | Tinnitus | Sensory Sensitivity | Tremors | Blurry Vision | Chest Pain | Menstrual | Twitching |
| --- | --- | --- | --- | --- | --- | --- | --- |
| 5 | 12 | 8 | 10 | 11 | 10 | 4 | 7 |

### Immune Profile Determined using a 14-Plex Cytokine/Chemokine Panel and Machine Learning

The 14-plex Cytokine/Chemokine panel was chosen from a total of 158 biomarkers because it identified a signature for PASC as previously published^10^. In order to determine whether these post-vaccination symptoms could be associated with similar 14 cytokine/chemokine abnormalities found in PASC, we performed machine learning on the 50 post-vaccination patients with PASC-like symptoms and compared the cytokine expression levels to individuals post-vaccination without PASC-like symptoms and healthy controls. The immune profile of post-vaccination individuals was classified as PASC using single classifier algorithm yet appeared to have less inflammation using a dual classifier (Figure 2) that included a severity index originally design to show the severity of disease in acute SARS-CoV-2.

Post-vaccination patients had statistically significant elevation in CCL5 (p=0.006), sCD40L (P<0.001), IL-8 (P<0.001), and IL-6 (P=0.04) (Figure. 3). Vaccinated individuals without PASC-like symptoms had elevated IL-2 (P=0.007) and reduced CCL4 (P=0.001) compared to the group of vaccinated individuals with PASC-like symptoms. Elevated IL-8 was a key difference in post-vaccine patients compared to the elevated cytokines in PASC.

**Figure 3.**
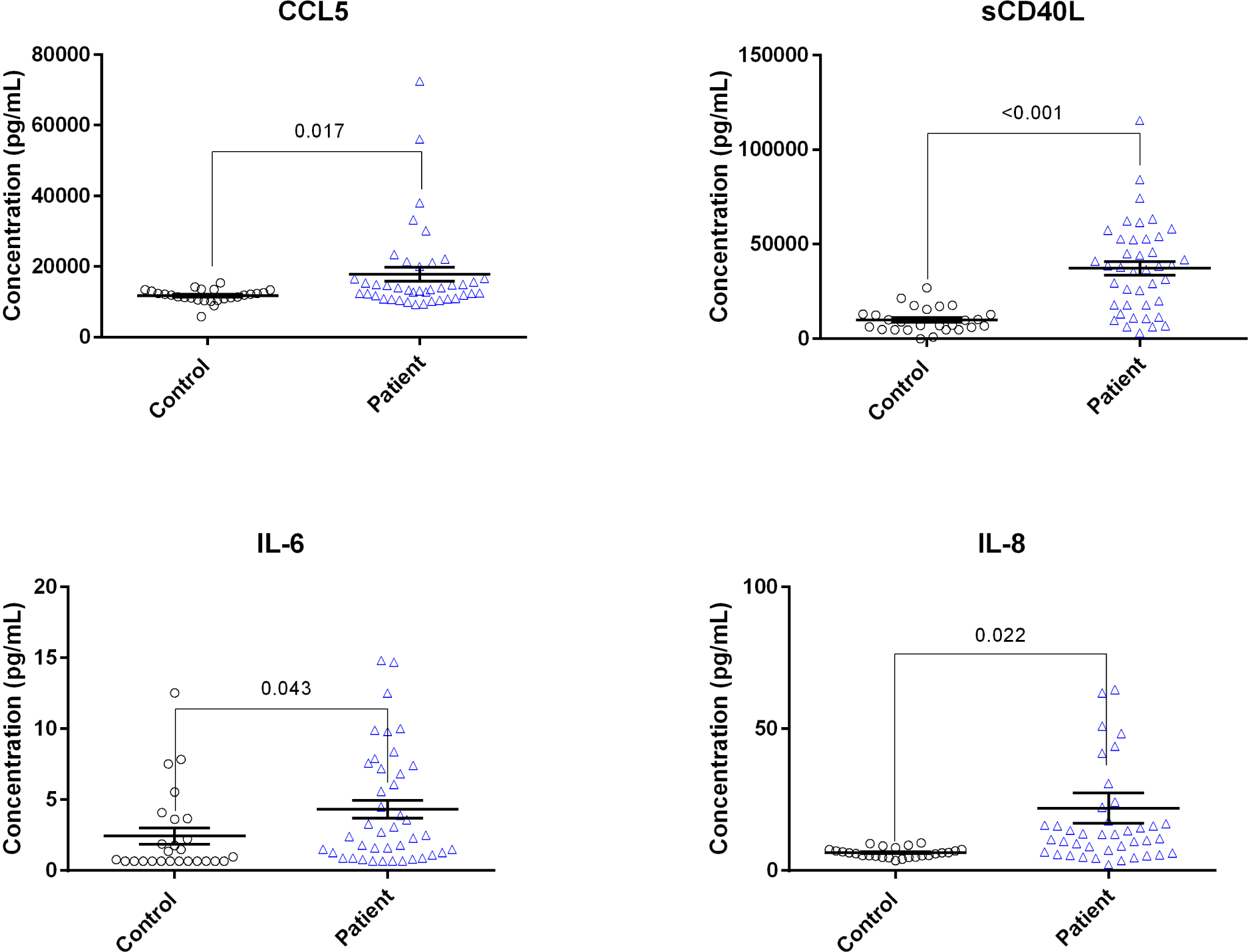
Statistically significant elevation of cytokines in post-vaccination individuals with PASC-like symptoms compared to vaccinated, healthy individuals.

### Analysis of S1 Protein Persistence

Similar to our published findings of S1 persistence in non-classical and intermediate monocytes in PASC^8^, we investigated whether a similar mechanism could account for the PASC-like symptoms in post-vaccination individuals with PASC-like symptoms. We used flow cytometry to screen patients for S1 protein in their monocyte subsets. We screened 14 post-vaccination individuals with PACS-like symptoms and 10 normal controls. As demonstrated in Figure 4, there was statistically significant elevation of S1 containing non-classical monocytes (13 of 14, P<0.001) and S1 containing intermediate monocytes (9/14, P=0.006).

**Figure 4.**
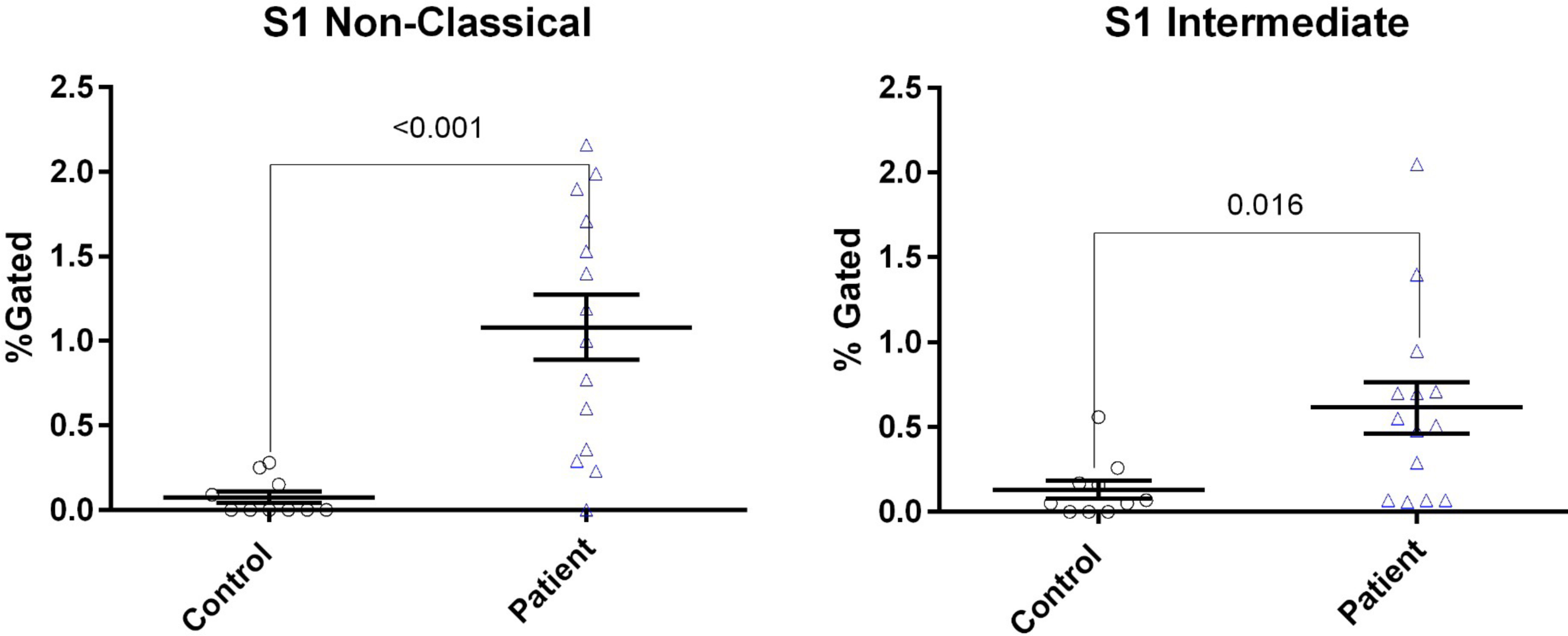
Flow cytometric quantification of S1-containing monocyte subsets as previously performed. Increased S1- conatining intermediate and non-classical monocytes was statistically significant compared to healthy controls.

Of the S1 positive post-vaccination patients, we sorted the CD16+ cells from six patients as previously performed for PASC patients^8^. Upon isolation of the protein, we demonstrated by LC-MS that these CD16+ cells from post-vaccination patients also contained S1 protein months after vaccination (Figure 5). Further analysis revealed that these CD16+ monocytes also contained peptide sequences of S2, and mutant S1 peptides.

**Figure 5.**
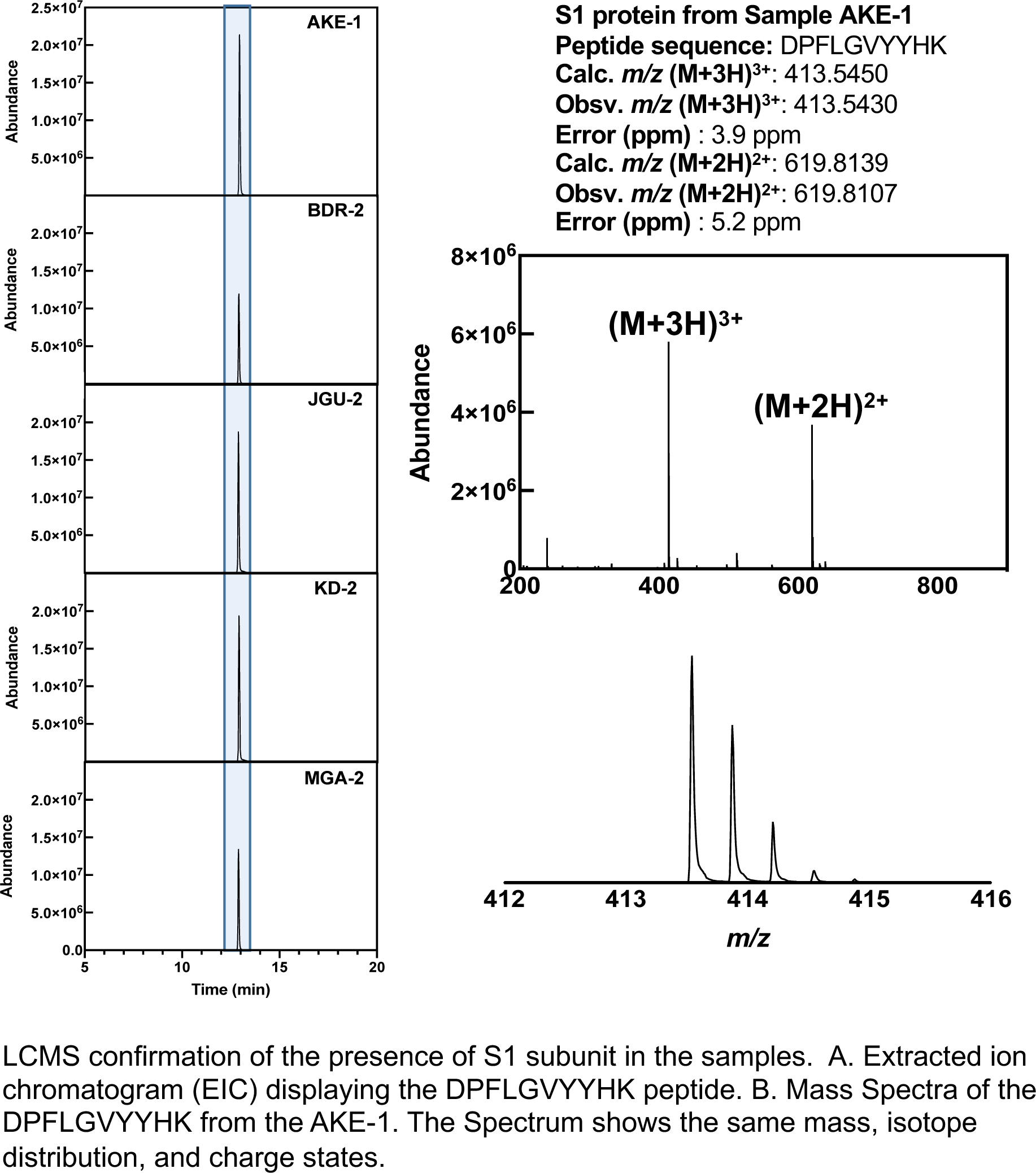
Liquid chromatography/mass spectrometry confirmation of S1 and S2 protein in flow cytometrically isolated cells from post-vaccination individuals with PASC-like symptoms

## DISCUSSION

To date, clinical manifestations of post-vaccination side effects or injuries have been described but very few mechanisms have been offered to explain these findings. In the present study, we investigated whether an S1 protein mechanism of inflammation similar to what we published in PASC might underlie the persistent, PASC-like symptoms that remain for months following vaccination with currently available vaccines in the United States and European Union.

Given that ongoing viral replication may not be required for prolonged symptoms^8^ and given the overlap in symptomatology, we applied machine learning to a panel of previously published immune biomarkers to determine if an immune signature for post-vaccination PASC-like symptoms might exist. Using two algorithms (severity score and long hauler index) previously derived from these biomarkers^10^, we found that post-vaccination PASC-like symptoms were associated with an inflammatory profile with statistically significant elevations in CCL5, sCD40L, IL-6, and IL-8. Further these patients were classified as PASC using a single classifier and PASC with inflammation using a dual classifier. Elevated IL-8 was a unique marker in post-vaccination individuals with PASC-like symptoms. We recently found a statistically significant correlation between decreased IL-8 and improvement in the NYHA cardiac symptom score in PASC following treatment with a CCR5 antagonist and statin^11^.

Because of the similarities between PASC and patients with post-vaccination PASC-like symptoms, we examined whether S1 protein persistence might also occur in patients with post-vaccination PASC-like symptoms. We demonstrated a statistically significant elevation of S1 protein containing non-classical monocytes (NCM) and in S1-containing intermediate monocytes (IM) in post-vaccination PASC-like patients compared to normal controls. We sorted these CD16+ monocytes as previously performed and used mass spectroscopy to interrogate whether S1 proteins were present in these highly mobile cells. We confirmed the presence of S1 sequences, S1 mutant amino acid sequences, as well as S2 sequences in the CD16+ monocytes from patients who represented all four vaccine manufacturers (Figure 6). Interestingly, we observed significance correlation (p=0.021) % S1 detected and days after vaccination in IM but no statistical significant correlation (p= 0.399) in NCM (Figure 7). In our previous study on the S1 protein persistence in PASC patients, these S1 mutant and S2 sequences were not detected^8^.

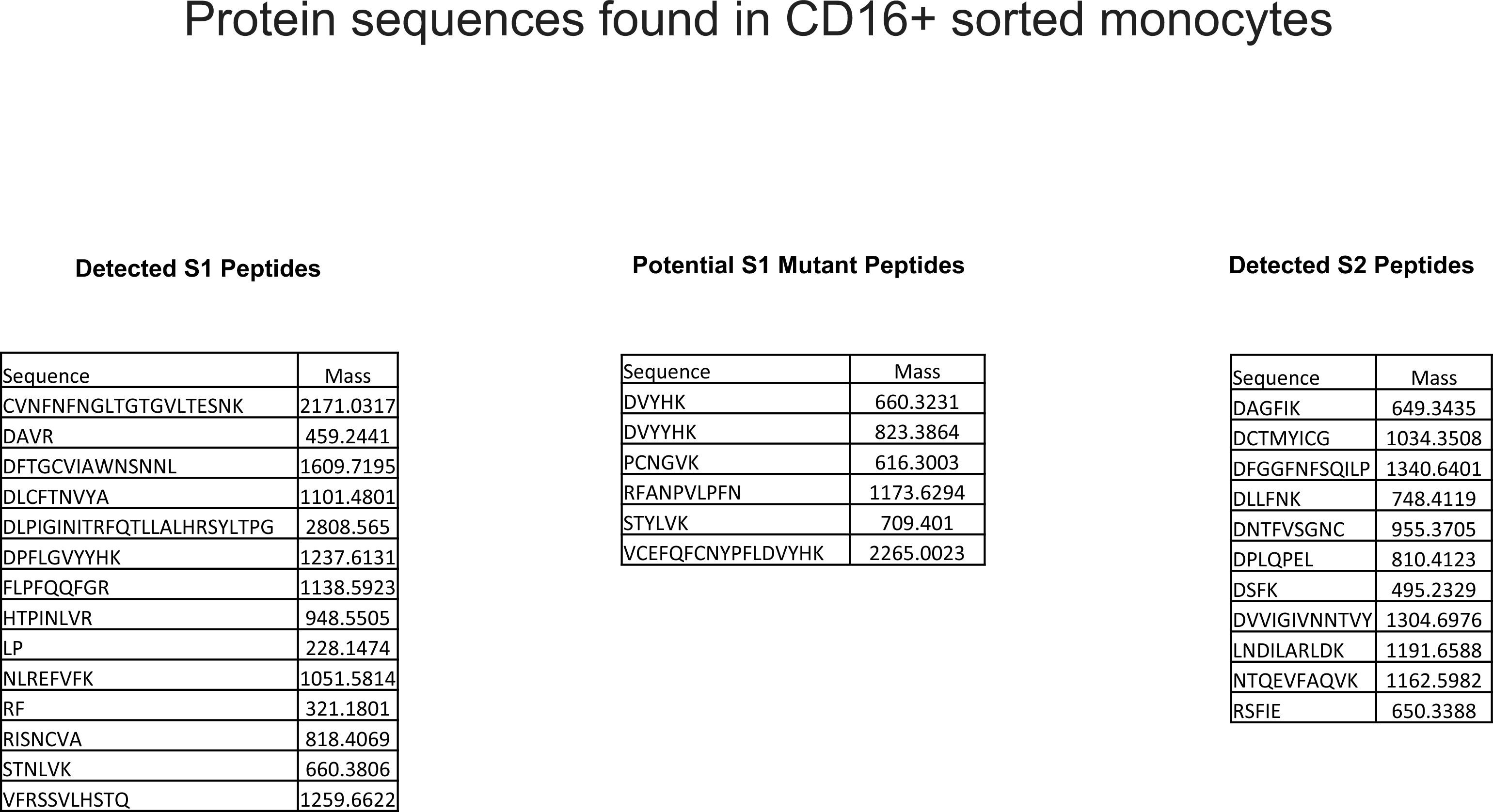

**Figure 7.**
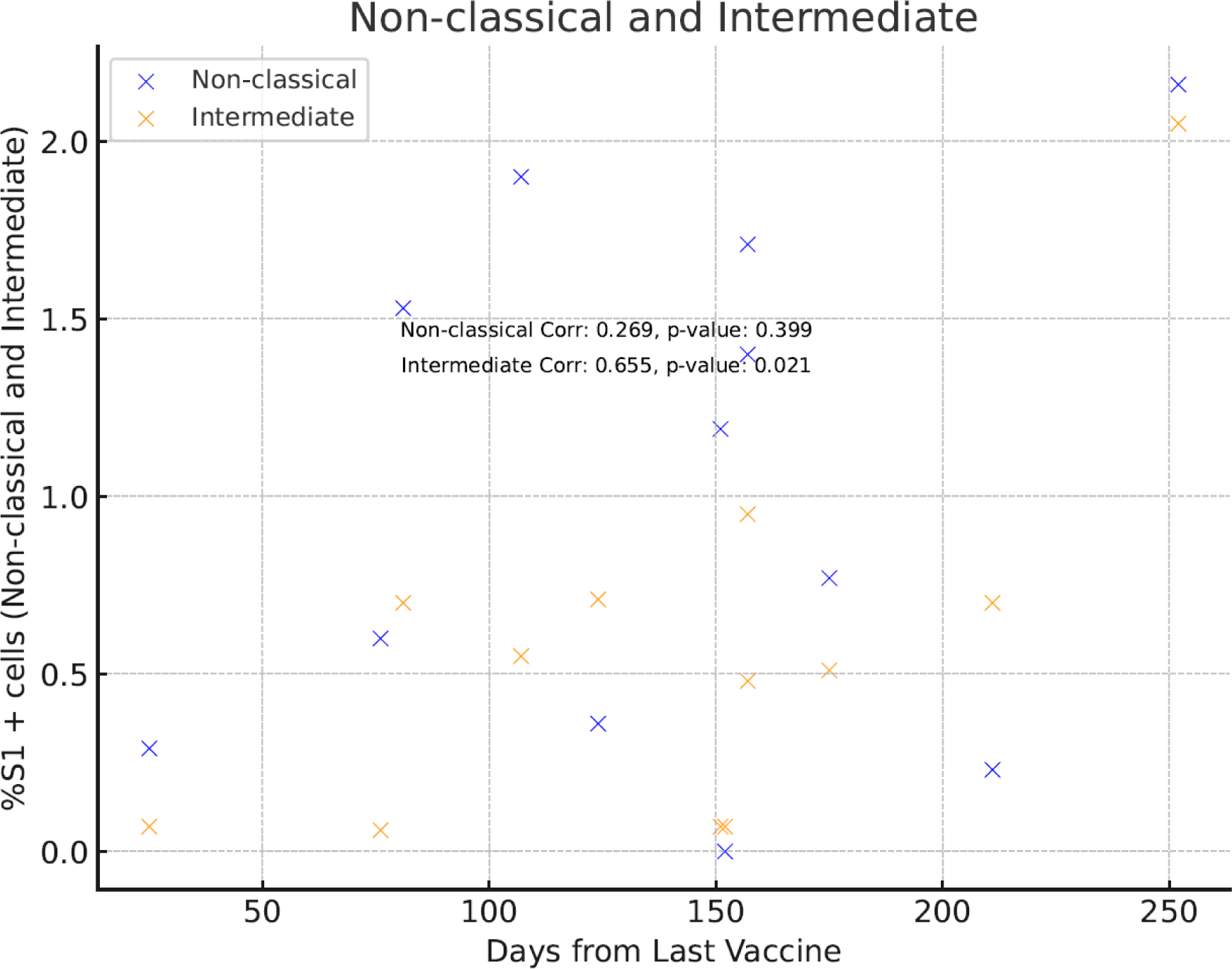
%S1 + cells vs the Days after the last COVID vaccination

Since the scope of this study was to also look for the presence of S1 proteins in CD16+ monocytes of post-vaccination patients, the clinical and pathological impact of the S1 mutant and S2 sequences remain unknown and will need to be elucidated through further studies.

NCM bind to fractalkine expressed on vascular endothelial cells through the expression of the fractalkine receptor CX3CR1 on the surface of NCM^12^. Fractalkine is also upregulated by IL-1, IFN-γ, and TNF-α^13^, cytokines that we have reported to be elevated in PASC^10^. CX3CR1 also provides a survival signal to non-classical monocytes through CX3CR1-dependent expression of the anti-apoptotic protein BCL2^14,15^. Stress and exercise mobilize non-classical monocytes including up to 4-fold with exercise^16,17^. The interaction between fractalkine and CX3CR1 has been involved in the pathogenesis of atherosclerosis, vasculitis, vasculopathies, and inflammatory brain disorders^18^ and could also be contributing to vascular endotheliitis in post-vaccination individuals with PASC-like symptoms. Vascular inflammation has been shown to expose the collagen surface and platelet activation/adherence by way of glycoprotein 1b-IX-V- receptor (GPIb-IX-V) with collagen-bound von Willebrand factor (vWF)^19,20^. Activated platelets also release soluble CD40 ligand (sCD40L) which leads to recruitment of both neutrophils and monocytes to the sites of vascular inflammation, thus activating the coagulation cascade^21^.

Activated platelets also release CCL5/RANTES which binds to endothelial cells, promoting monocyte adhesion to inflamed endothelial tissues^22^ and acting as a chemoattractant for inflammatory cells. Studies in atherosclerosis have shown that CCR5 antagonists reduce non-classical monocyte recruitment to sites of atherosclerosis^22,23^. In addition, accumulation of non-classical monocytes can be reduced by statin treatment through reduction in fractalkine expression^24,25^. Interfering with these pathways may hold potential therapeutic targets for PASC and post-vaccination individuals with PASC-like symptoms.

Further, activated platelets and endothelial cells also secrete (VEGF) which induces angiogenesis and microvascular hyperpermeability. VEGF contributes to vasculitic neuropathy and also promotes a pro-inflammatory-prothrombotic environment^26^. This pathway may also contribute to the thrombosis seen in some individuals post-vaccination.

Taken together, these findings suggest a possible mechanism for the debilitating symptoms found in some patients weeks and months following vaccination. The findings that the immune profile and persistent S1 protein in CD16+ monocytes suggest that S1 protein persistence is a major contributor not only of symptoms in PASC but also may be a major contributor of persistent post COVID vaccination complications itself given that the S1 delivered by vaccination and thus absence of viral replication can cause similar pathologic features.

A significant limitation of this study was the lack of approved testing to 100% rule out previous infection and it is possible the persistent S1 protein detected in the CD16+ monocytes of some of the patients in this study is from SARS-CoV-2 and not from the vaccine. There also exists the possibility that some of these new-onset symptoms post-COVID vaccination are unrelated to the vaccines. The data from this study also cannot make any inferences on epidemiology and prevalence for persistent post-vaccine symptoms. Thus, further studies and research need to be done to understand the risk factors, likelihood and prevalence of these symptoms.

## Ethics Statement

The independent Chronic COVID Treatment Center (CCTC) Ethics and IRB group reviewed and approved the study. All the patients/participants provided their written informed consent to participate in this study.

## Funding

None

## Author contributions

BP and RY conceptualized the study. RY organized the study. JG-C,and RM-R performed the bioinformatics. All authors contributed to revising the manuscript and approved the submitted version.

BP, RY, JM, EBF, JGC, RM-R wrote the draft of the manuscript and all the authors contributed to revising the manuscript prior to submission.

## Competing Interests

BP, EBF, EL, CB, and AP are employees of IncellDX.

BP, RY, JB, EO, DJ, and MK are independent contractors of the CCTC.

## Data and materials availability

All requests for materials and raw data should be addressed to the corresponding author

## Data Availability

All data produced in the present study are available upon reasonable request to the authors

---

| Patient | Vax | NCM Pat | Int Pat | Days from<br>Last Vaccine |
| --- | --- | --- | --- | --- |
| 1 | PFE | 1.19 | 0.07 | 151 |
| 2 | MRNA | 1.71 | 0.48 | 157 |
| 3 | ASTRA | 2.16 | 2.05 | 252 |
| 4 | MRNA | 0.6 | 0.06 | 76 |
| 5 | MRNA | 1.9 | 0.55 | 107 |
| 6 | PFE | 0.36 | 0.71 | 124 |
| 7 | PFE | 0.29 | 0.07 | 25 |
| 8 | PFE | 0.23 | 0.7 | 211 |
| 9 | PFE | 1.53 | 0.7 | 81 |
| 10 | MRNA | 1.4 | 0.95 | 157 |
| 11 | PFE | 0.77 | 0.51 | 175 |
| 12 | PFE | 0 | 0.07 | 152 |

## Notes

### Funding Statement

This study did not receive any funding

### Author Declarations

Ethics Statement: The independent Chronic COVID Treatment Center (CCTC) Ethics and IRB group reviewed and approved the study. All the patients/participants provided their written informed consent to participate in this study.

